# A Study Analysing the Distribution and Determinants of Diabetic Macular Edema in a Tertiary Care Center

**DOI:** 10.1101/2024.11.23.24317493

**Authors:** M. S. Priethikka, B. Chandrasekaran, L Subha, Vikram Chellakumar, M Balakrishnan, Deepthi Solasa

**Affiliations:** Junior Resident, Dept. of Ophthalmology, Sree Balaji Medical College and Hospital; Professor & Head of Dept., Dept. of Ophthalmology, Sree Balaji Medical College and Hospital; Associate Professor, Dept. of Ophthalmology, Sree Balaji Medical College and Hospital; Professor, Dept. of Ophthalmology, Sree Balaji Medical College and Hospital; Senior Resident, Dept. of Ophthalmology Sree Balaji Medical College and Hospital

## Abstract

**AIM:** This study aims to examine the correlation between the specific type of diabetic macular edema (DME) identified using Optical Coherence Tomography (OCT) and various factors, including patient age, gender, diabetic profile (fasting blood sugars, postprandial blood sugars, and HbA1c), duration of Type 2 Diabetes Mellitus, and central macular thickness on OCT.

**OBJECTIVE:** The study intends to investigate the relationships between the age of patients and DME type, gender prevalence in DME, duration of Type 2 Diabetes Mellitus and DME type, diabetic profiles and DME type, central macular thickness and DME type, and severity of diabetic retinopathy and DME type.

**Introduction:** Diabetic maculopathy is a major cause of vision impairment in diabetic retinopathy. This study explores the relationship between DME types as determined by OCT and factors such as age, gender, diabetic profile, and diabetes duration.

**Materials and Methods:** Conducted over one year at a tertiary health care center, the study evaluated 95 patients with diabetic maculopathy through comprehensive clinical assessments including OCT classification of DME types.

**Results:** The findings indicated a higher prevalence of DME among males, predominantly in the 61- 70 age group. A significant association was found between diabetes duration and mixed-type DME. However, no significant correlations were observed between glycemic control measures (FBS, PPBS, HbA1c) and DME types.

**Conclusion:** The study underscores the importance of demographic and clinical factors in understanding variations in DME types, highlighting the need for tailored management strategies to reduce vision loss risk in diabetic patients.

## INTRODUCTION

Diabetes mellitus, comprises a group of prevalent endocrine disorders characterized by consistently elevated blood sugar levels. This condition arises from either insufficient insulin production by pancreas or decreased responsiveness of body’s cells to insulin. Neglected, diabetes can precipitate various health complications affecting the cardiovascular system, eyes, kidneys, and nerves, contributing to approximately 1.5 million annual deaths.

Diabetic retinopathy is a common complication of diabetes, affecting approximately one-third of adults diagnosed with the disease. It is a leading cause of vision loss in middle-aged and elderly population and occurs when high blood sugar levels damage the blood vessels in the retina.

Diabetic retinopathy typically progresses through several stages:

□ Non-proliferative diabetic retinopathy: This early stage is characterized by weakened blood vessels in the retina, leading to microaneurysms, hemorrhages, and fluid leakage.
□ Proliferative diabetic retinopathy: In this advanced stage, the retina receives insufficient blood supply, triggering the growth of new, abnormal blood vessels. These vessels are fragile and prone to bleeding, potentially causing severe vision loss or blindness.

Macular thickening due to diabetic retinopathy stems from retinal vascular hyperpermeability and microenvironmental changes, often causing vision loss in individuals with diabetes. It can manifest across a spectrum of retinopathy severity, from mild to severe. Areas of retinal vascular incompetence, visualized by angiography, as well as regions of retinal ischemia, can contribute to Diabetic Retinopathy development. This thickening may arise from various factors, including leakage from microaneurysms or hyperpermeable capillaries, and ischemia-induced swelling.

Optical Coherence Tomography plays a crucial role in classification and management of diabetic macular edema, a common complication of diabetic retinopathy that can lead to vision impairment if not properly managed. OCT provides detailed cross-sectional imaging of the retina, allowing clinicians to visualize and classify various types of macular edema based on their characteristics.

OCT is used to monitor the response to various treatments for DME, such as intravitreal anti-VEGF injections, corticosteroid implants or laser treatments. Reduction in central subfield thickness and resolution of fluid on OCT scans indicate treatment efficacy.

## REVIEW OF LITERATURE

### DIABETES MELLITUS

Diabetes is a complex, chronic condition requiring continuous medical care with multifactorial risk-reduction strategies beyond glycemic management (1). Classic symptoms include increased thirst, frequent urination, weight loss, and vision disturbances (2). Untreated, diabetes can lead to various micro and macrovascular complications (3). These include macrovascular complications like coronary heart disease, stroke, and peripheral arterial disease, as well as microvascular complications such as diabetic kidney disease, retinopathy, and peripheral neuropathy. Heart failure is often a common manifestation of cardiovascular disease in people with type 2 diabetes, increasing the risk of death in those with type 1 or type 2 diabetes (5). While these complications still pose significant health challenges, their occurrence is decreasing due to better diabetes management.

However, as individuals with diabetes now live longer, they are encountering a new range of health issues. Advances in diabetes care have shifted focus towards conditions like cancer, infections, functional and cognitive impairments, liver disease, and affective disorders, which are becoming increasingly associated with diabetes(6).

Other specific types include diabetes caused by genetic defects such as maturity-onset diabetes of the young (MODY) and other rare genetic disorders leading to neonatal diabetes, as well as conditions affecting the pancreas. Gestational diabetes during pregnancy is also commonly observed. The clinical presentation of these genetic and secondary forms of diabetes typically resembles that of type 2 diabetes, with severity influenced by the extent of beta cell dysfunction and insulin resistance present(7).

### EPIDEMOLOGY

Approximately 422 million individuals globally have diabetes, with majority residing in low- and middle-income nations. Each year, diabetes is directly responsible for 1.5 million deaths. Over recent decades, both the number of cases and the prevalence of diabetes have been consistently rising (8).

In 2014, 8.5% of adults aged 18 years and older were affected by diabetes. By 2019, diabetes directly caused 1.5 million deaths, with 48% occurring before the age of 70. Additionally, diabetes contributed to 460,000 deaths from kidney disease, and about 20% of cardiovascular deaths were attributed to elevated blood glucose levels (9).

In contrast, globally, the likelihood of premature death (between ages 30 and 70) from any of the four major non-communicable diseases (cardiovascular diseases, cancer, chronic respiratory diseases, or diabetes) decreased by 22% between 2000 and 2019 (WHO).

According to the ICMR, INDIA B study done between 2022- 2023 the overall weighted prevalence of diabetes in India was 11.4%.

As of 2021, an estimated 537 million individuals worldwide had diabetes, representing 10.5% of the adult population, with type 2 constituting about 90% of cases (1). Projections suggest that by 2045, approximately 783 million adults will have diabetes, marking a 46% increase from current statistics.

The traditional complications of diabetes mellitus are well known and continue to pose a considerable burden on millions of people living with diabetes. (10). According to ICMR, the prevalence of chronic diabetes complications ranges from 4.8-21.7% for retinopathy, 0.9-62.3% for nephropathy and 10.5-44.9% for neuropathy (11).

Regarding the epidemiology of diabetic retinopathy, the prevalence at the time of diagnosis differs between type 1 and type 2 diabetes. In type 1 diabetes, the prevalence ranges from 0- 3% (12), whereas in type 2 diabetes, the prevalence is higher. Prevalence of DR in India ranges from 17.6% to 28.2% (13)(14)(15) and less than 10% of the persons with diabetes suffer from sight-threatening DR which includes diabetic macular edema and proliferative diabetic retinopathy (13). Diabetic macular edema (DME), swelling in the macula caused by fluid leaking from retinal blood vessels, can occur with any stage of diabetic retinopathy. Diabetic retinopathy, VTDR, and DME affect 28.5%, 4.4%, and 3.8%, respectively of adults 40 years and older with diabetes (16) (17).

### DIABETIC RETINOPATHY

Diabetic retinopathy (DR) is a progressive eye condition that primarily affects the retinal blood vessels in individuals with diabetes. Retinopathy is already present at the time of diagnosis in 20% of patients with type 2 diabetes (18). It can be categorized into two main stages: non-proliferative diabetic retinopathy (NPDR) and proliferative diabetic retinopathy (PDR).

Diabetic retinopathy is characterized by increased permeability of retinal blood vessels, leading to fluid accumulation in retina. This stage also involves capillary closure, resulting in the formation of microaneurysms, venous dilation, retinal hemorrhages, and hard exudate deposition (19), (20). As the disease progresses to proliferative diabetic retinopathy, retinal ischemia caused by capillary occlusion triggers neovascularization – the growth of fragile new blood vessels that are prone to hemorrhage, potentially causing vitreous hemorrhage and tractional retinal detachment, which can lead to severe vision loss (21).

To assess the severity of diabetic retinopathy, various scales, have been developed, including the International Clinical Diabetic Retinopathy Disease Severity and Diabetic Macular Edema Severity Scales, which build upon earlier research-oriented scales such as those from the Early Treatment Diabetic Retinopathy Study and the Wisconsin Epidemiologic Study of Diabetic Retinopathy (22). Regular eye examinations and proper management of diabetes are crucial for early detection and treatment of diabetic retinopathy, as it may not present symptoms in its early stages (19).

### BIOCHEMICAL PATHWAYS LINKING HYPERGLYCEMIA TO VASCULAR DAMAGE

The pathogenesis of vascular damage and the subsequent development of macular edema is a complex process that involves a diverse array of biochemical pathways, all of which are collectively influenced by hyperglycemia. These pathways include metabolic changes, hemodynamic alterations, leukostasis, mechanical or tractional forces from vitreous body, and influence of angiogenic growth factors (23).

There are multiple metabolic pathways involved in the pathogenesis such as the Sorbitol pathway, Protein C kinase pathway, Diacylglycerol pathway, formation of advanced glycation end product pathway and vascular endothelial growth factor pathway (24).

### SORBITAL PATHWAY

One of the significant metabolic alterations induced by hyperglycemia is the activation of the sorbitol-polyol pathway (25),(26). In tissues with low insulin dependence, such as neurons, glomeruli, lens and retina, elevated glucose levels lead to increased intracellular glucose. In this process, the enzyme aldose reductase catalyzes the conversion of glucose to sorbitol, which is then further converted to fructose by sorbitol dehydrogenase, with NAD+serving as a cofactor. Sorbitol, unable to easily traverse cell membranes, accumulates within cells, potentially causing cellular damage through osmotic effects, particularly in tissues like the lens (25), (27).

Additionally, sorbitol accumulation can lead to increased NADH/NAD+ ratios, a condition referred to as pseudohypoxia; and depletion of intracellular myoinositol. Myoinositol, which is structurally similar to glucose and serves as a precursor to phosphatidylinositol, plays a crucial role in activating Na+-K+- ATPase directly or through diacylglycerol (28).

### PROTEIN KINASE C PATHWAY

Pseudohypoxia affects lipid metabolism, enhances free radical production, and triggers the activation of protein kinase C isoforms. The increased oxidative stress and inflammatory processes contribute to the impairment of the blood-retinal barrier, which is a crucial factor in the development of diabetic macular edema(29), (30), (31).

Antioxidants, on the other hand, have been found to hinder sorbitol accumulation, advanced glycation end-product formation, and Protein C kinase activation, all of which are implicated in the pathogenesis of diabetic microvascular complications (32), (33). Tocopherol, a type of antioxidant, has been observed to prevent diacylglycerol and protein kinase C activation, as well as the hemodynamic abnormalities associated with diabetic retinopathy in animal models (34), (35).

### CLINICALLY SIGNIFICANT MACULAR EDEMA

Clinically significant macular edema (CSME) is a condition characterized by visual impairment resulting from the presence of fluid accumulation in the macula.

According to the Early Treatment Diabetic Retinopathy Study Research Group, clinically significant macular edema is defined by retinal thickening within 500 micrometers of the fovea, the presence of hard exudates at or within 500 micrometers of the fovea with adjacent retinal thickening, or a zone of retinal thickening one disc area or larger, part of which is within one disc diameter of the fovea (36), (37), (38). According to the International classification of diabetic retinopathy, Diabetic macular edema is classified into,

Center-involved DME: Retinal thickening in the macula that involves the central subfield zone (1 mm in diameter)

Non-center involved DME : Retinal thickening in the macula that does not involve the central subfield zone (1 mm in diameter)

### PATHOGENESIS OF DIABETIC MACULAR EDEMA

Macular edema is characterized by an anomalous accumulation of fluid in the macula. Extracellular fluid can infiltrate retinal layers, accumulate in cavities commonly referred to as “cysts” (39).

The exact mechanisms contributing to DME include:

1. Blood-Retinal Barrier (BRB) Dysfunction: Chronic hyperglycemia leads to breakdown of the blood retinal barrier, which consists of tight junctions between retinal endothelial cells, allowing fluid, lipids, and proteins to leak into the macula (39) (40).
2. Increased Vascular Permeability: Elevated levels of vascular endothelial growth factor and other inflammatory mediators contribute to increased permeability of retinal capillaries, exacerbating fluid leakage into the macula (39).
3. Ischemia and Hypoxia: It occurs when there is an inadequate blood flow and oxygen delivery to the retina as a result of diabetic microvascular changes leading to tissue hypoxia, further stimulating VEGF production and exacerbating edema(39).

### OPTICAL COHERENCE TOMOGRAPHY IN DIABETIC MACULAR EDEMA

OCT is a diagnostic technology that provides a non-invasive and real-time cross-sectional image of the retina. OCT employs. interferometry to generate a precise cross-sectional depiction of the retina, with an accuracy of at least 10-15 microns (41). OCT was first introduced in 1991 by Huang and colleagues.

OCT provides a detailed analysis of fluid accumulation patterns in Diabetic macular edema. Various studies have documented diverse patterns of fluid distribution in diabetic macular edema, such as Diffuse retinal thickening, Cystoid macular edema, Serous retinal detachment, and Posterior hyaloid traction (42).

OCT has identified several qualitative biomarkers that have prognostic value for visual outcomes in diabetic macular edema, such as Disorganization of retinal inner layers, Integrity of the Ellipsoid zone, and presence of Hyper-reflective foci.

Fluid Accumulation patterns: OCT distinguishes between different patterns of fluid accumulation within the retina, which are crucial for guiding treatment choices:

- Cystoid Macular Edema : Characterized by the presence of cystic spaces within the macula, typically seen as round or oval hyporeflective areas on OCT scans (43).
- Diffuse Retinal Thickening: Involves generalized thickening of the retina without distinct cystoid spaces (43).
- Serous Retinal Detachment: Refers to the accumulation of fluid between the neurosensory retina and retinal pigment epithelium, appearing as a hyporeflective space beneath the retina on OCT images (43).
- Vitreo-macular traction : The pathological basis for these clinical observations is that in diabetes, the vitreous undergoes abnormal glycation of its collagen and other structural components, leading to increased stiffness and the potential for traction on the retina (44).
- Mixed type: It is a composite of multiple types of DME (45).

### ADDITIONAL VARIABLES

Central Subfield Thickness: OCT measures the thickness of the central subfield, which is a key parameter in assessing the severity of diabetic macular edema. Elevated CST is indicative of macular edema and provides a quantitative measure of the degree of retinal thickening (45), (46).

Subretinal Fluid : It refers to the fluid accumulation between the neurosensory retina and the retinal pigment epithelium, often associated with leakage from choroidal vessels or exudation from retinal vessels (47).

### OCT BIOMARKERS

Quantitative Biomarkers

□ Central Retinal Thickness: Objective measurement of retinal thickness that can be correlated with visual acuity.
□ Cube Average Thickness and Cube Volume: Additional quantitative measures of retinal thickness and volume.

Qualitative Biomarkers

□ Subretinal Fluid (SRF): Presence of fluid accumulation beneath the retina.
□ Intraretinal Cysts (IRC): Fluid-filled cysts within the retinal layers.
□ Ellipsoid Zone Disruption : Disruption of the ellipsoid zone, a marker of photoreceptor integrity (48).
□ Disorganization of Retinal Inner Layers : Disruption of the normal retinal layer architecture (48) (49).
□ Hyperreflective Foci: Tiny hyperreflective dots within the retina, associated with inflammation (50).
□ Epiretinal Membrane: Fibro-cellular proliferation on the inner retinal surface (51).
□ Vitreomacular Interface Changes: Alterations in the vitreoretinal interface (52).
□ Hard Exudates: Lipid deposits within the retina.

These OCT biomarkers provide valuable information about the structural and functional changes in the retina associated with DME. They can help guide treatment decisions and predict visual and anatomical outcomes in DME patients.

- Cystoid macular edema
- Diffuse Spongiform retinal thickening
- Serous macular detachment
- Mixed type
- Taut posterior hyaloid

Treatment of Diabetic Retinopathy:

Diabetic retinopathy is a severe microvascular complication of diabetes mellitus that can lead to vision loss if left untreated. The evidence-based management of diabetic retinopathy involves a two-pronged approach: indirect management through optimization of glycemic control and systemic hypertension, and direct management of the ocular condition.

Optimizing glycemic control and systemic hypertension is crucial in slowing the onset and progression of diabetic retinopathy.Two landmark clinical trials, the Diabetes Control and Complications Trial and the United Kingdom Prospective Diabetes Study, have conclusively demonstrated that intensive glycemic control significantly reduces the risk of developing and progressing diabetic retinopathy in both type 1 and type 2 diabetes. The DCCT study showed that intensive glycemic control resulted in a 76% reduction in the onset of diabetic retinopathy, a 63% reduction in its progression, and a 47% reduction in the development of proliferative diabetic retinopathy (53).

Intravitreal injections (IVI) of anti-VEGF agents are the preferred initial treatment for newly diagnosed center-involved diabetic macular edema (CI-DME). Several clinical trials have proven that anti-VEGF therapy is more effective than traditional macular laser treatment for improving vision in CI-DME, establishing it as the primary therapy.

The Diabetic Retinopathy Clinical Research Network protocol for CI-DME typically begins with monthly IVI for 4–6 months initially (54). Subsequently, treatment may be paused if there is no improvement in vision or central subfield thickness, or if significant visual acuity (6/6 on Snellen chart) or resolution of DME is achieved. If consecutive visits show no need for treatment, the interval between visits may be extended up to 4 months, which has been shown to reduce the number of injections while maintaining good visual acuity gains (55).

Treatment can be deferred once vision improves and optical coherence tomography shows improved findings. An alternative strategy to lessen injection frequency is the treat-and-extend regimen, where the interval between visits is adjusted based on the patient response to treatment. Recent trials have demonstrated that this approach achieves comparable visual and anatomical outcomes over two years compared to monthly dosing, with fewer injections required (56).

Laser photocoagulation is one of the direct treatment modalities for diabetic retinopathy. Studies such as the Diabetic Retinopathy Study and the Early Treatment Diabetic Retinopathy Study have demonstrated the efficacy of laser treatment in reducing the risk of severe vision loss in patients with proliferative diabetic retinopathy and clinically significant macular edema (57). However, in advanced stages of the disease, treatment options may be limited, and vitreo-retinal surgery may be required.

## METHODOLOGY

The investigation was conducted at a tertiary multispeciality Hospital over a period of one year, and it involved both inpatients and outpatients. A total of 95 patients diagnosed with Diabetic Maculopathy were evaluated. The OCT findings were the primary focus in order to ascertain the type of maculopathy. A comprehensive medical history was obtained, which encompassed the age, gender, duration of diabetes, the degree of diabetes control, and any other co-existing medical conditions. The Snellen chart was employed to ascertain the best corrected visual acuity that could be attained. Goldman applanation tonometry was employed to measure the intra-ocular pressure. A slit lamp was employed to evaluate the anterior segment. Macular functionality was assessed using the Amsler grid. Zeiss 4 mirror gonioscopy lens was employed to assess the angle.

The pupil was dilated by administering eye drops containing 10% phenylephrine and 1% tropicamide. Direct ophthalmoscopy and Indirect ophthalmoscopy, as well as slit lamp examination with a +90D lens, were employed to examine the fundus. Additionally, the fundus was meticulously examined and documented in other regions using an Indirect ophthalmoscope in conjunction with a +20D lens. The fundus image was captured using a Zeiss fundus picture camera. SD- OCT, was implemented to evaluate the macula. This enabled the identification and classification of the type of Diabetic Retinopathy and the specific form of Diabetic macular edema. Patterns and connections were identified through the recording and analysis of all observations.

### INCLUSION CRITERIA

- All age group with type 2 Diabetes Mellitus.
- Patients who have Diabetic retinopathy associated with any form of Diabetic macular edema involving the macula.

### EXCLUSION CRITERIA -

- Patients with diabetic retinopathy with no macular involvement.
- Other retinal vascular disease.
- Glaucoma.
- Uveitis.
- Grade 3 and 4 Hypertensive retinopathy.

## RESULTS

In this study, the most affected age group was 61-70 years, followed by 51-60 years, 71-80 years, and 41-50 years. This corresponds with the findings of the Wisconsin Epidemiologic Study of Diabetic Retinopathy, which showed a higher prevalence of diabetic retinopathy in individuals in the middle aged and elderly population. The study evaluated age in relation to the type of diabetic macular edema but did not find statistically significant results. Therefore, age may not be associated with the type of macular edema.

Out of the 95 patients with diabetic macular edema, 66% were male and 34% were female. The relationship between gender and the type of diabetic macular edema was assessed, but no statistically significant p values were found. This suggests that gender may not be associated with the type of macular edema.

Twelve patients who had diabetes mellitus for 11 to 20 years experienced a mixed type and cystoid type of diabetic macular edema more frequently. A similar trend was observed in patients with diabetes duration of 21 to 30 years. This suggests that in diabetes of longer duration, mixed type and cystoid type of DME are predominant. The statistical significance of these findings is supported by a significant p value.

Out of the 95 patients, the highest incidence of diabetic macular edema (DME) was observed in severe non-proliferative diabetic retinopathy (NPDR). Patients with proliferative diabetic retinopathy typically exhibited mixed-type DME, while those with severe NPDR tended to present with cystoid-type DME. These findings showed statistically significant values.

The study examined fasting blood sugar (FBS) postprandial blood sugar (PPBS) and glycated hemoglobin in relation to the type of diabetic macular edema but did not find any statistically significant associations. This suggests that FBS, PPBS, HBA1C levels may not be correlated with the type of macular edema. However, people with higher level of glycated hemoglobin have higher chances of developing Diabetic retinopathy.

Furthermore, patients showed diverse levels of glycemic control influenced by their medication regimens—either oral or insulin—and varied durations of diabetes. Thereby, blood sugar levels at the time of presentation could not be accurately assessed.

## DISCUSSION

This study was carried out in a tertiary care multispeciality hospital. It involved 95 patients diagnosed with Diabetic macular edema, without any specific age restrictions. The purpose of our study was to investigate the correlation between different demographic, clinical, and OCT parameters and the OCT classification of Diabetic Macular Edema.

This study demonstrates that within the entire screened sample population, there was a higher proportion of males compared to females. (58). According to a decade-long cross-sectional study conducted by Lundeen et al., there were variations between men and women in the yearly occurrence rates of DME or VTDR, with men showing a higher prevalence compared to women. However, these results were not statistically significant in our study.

In this study the predominant age group affected was between 61-70 years followed by 51-60 years and then 71 - 80 years range and 41 – 50 years range and diabetic macular edema along with diabetic retinopathy was more commonly seen in the middle age or elderly population.

This correlates with the Winconsin Epidemiologic study of Diabetic Retinopathy revealing that diabetic retinopathy was more prevalent in the middle aged and elderly population affecting above 50 years of age. (22) Firstly, the duration of diabetes is typically longer in older patients, and the prevalence of diabetic retinopathy is strongly associated with the duration of the disease. Additionally, older patients may have a higher risk of developing other comorbidities, such as hypertension and cardiovascular disease, which can also contribute to the development and progression of diabetic retinopathy (59).

The duration of diabetes when seen in relation to the type of diabetic macular edema showed that longer the duration of diabetes and its related changes, mixed type of DME was seen followed by cystoid type of DME. These results showed statistical significance in our study. A meta-analytic study conducted by Ryan Lee and Tien Y Wong analyzed 35 studies conducted between 1980 and 2008. The review indicated that the incidence of DR among individuals with diabetes was 35.4%. Additionally, the prevalence of DME in this cohort ranged from 2% to 12.3% (59), (60).

Additionally, according to a study done by Klein R, Knudtson M, Lee K, Gangnon R et al., they assert that both DR and DME have numerous shared risk factors including duration of diabetes, pregnancy, hypertension, hyperglycemia, dyslipidemia and obesity. They analysed that the duration was a significant risk factor for DR, and this is independent of adequacy of glycemic control (61).

Our study investigated the correlation between glycemic profile (FBS,PPBS,HBA1C) of the patient and the type of diabetic macular edema on OCT and no significance was found. While other studies have also explored the potential link between glycemic control and the development of diabetic macular edema, the results have been inconclusive. A study conducted by Cheung et al., evaluated the relationship between fasting blood sugar (FBS), postprandial blood sugar (PPBS) and HBA1C with the type of diabetic macular edema, but found no significant association (26).

When comparing the type of Diabetic retinopathy with the type of Diabetic macular edema, it is important to note that DME can occur in patients at any stage of DR. However, the prevalence of DME tends to increase as the severity of clinical DR grading (62). Our study revealed that severe non-proliferative diabetic retinopathy (NPDR) was more frequently observed, and patients with proliferative diabetic retinopathy often exhibited vitreomacular traction on OCT imaging.

The central macular thickness when evaluated with the type of Diabetic macular edema proved that increase in thickness correlated significantly with mixed type of DME followed by cystoid type of DME (63). As longer duration very commonly presents with more than one type of presentation on OCT. Qi Sheng You, MD, PhD; Kotaro Tsuboi conducted a cross sectional observational study on 215 patients with diabetes where they compared the relationship between Central macular thickness and diabetic macular edema with the help of OCT-A. The study found a significant correlation between DME and CMT, with a correlation coefficient of −0.303 (P&lt; .001) (63), (64) .

The frequency and intensity of diabetic retinopathy, which includes retinal edema, are strongly associated with management of blood sugar levels. Implementing stricter regulation of blood glucose levels and reducing blood pressure can effectively decrease the likelihood of diabetic retinopathy advancing. Nevertheless, the precise processes via which hyperglycemia leads to the formation of diabetic macular edema remain incompletely comprehended.

The patients in our study predominantly underwent Anti-VEGF treatment according to the DRCR.net protocol. Individuals with Proliferative diabetic retinopathy underwent an initial procedure called Pan-retinal photocoagulation, followed by further therapy with Anti-VEGF as needed. Patients who exhibited increased levels of inflammatory biomarkers on OCT were administered intravitreal steroids after receiving three doses of Anti VEGF, as deemed necessary. The majority of patients with vitreomacular traction or adhesions received surgical management.

## LIMITATIONS

1. Sample size being 95 only concentrated on the population attending this hospital.
2. This being a Cross sectional study had no follow up to understand the course of the type of DME.
3. Patients were diversified with different medical and treatment histories thereby limiting our understanding of the condition.

## CONCLUSION

The study indicated that diabetic retinopathy and diabetic macular edema were more prevalent in middle-aged and elderly individuals. It was found that the type of DME observed through optical coherence tomography did not significantly correlate with the patient’s age. DME was more frequently observed in males compared to females, although the classification of DME on OCT was not affected by gender. Additionally, the laterality of the eye had no significant impact on the type of DME identified on OCT. A longer duration of diabetes and associated ocular changes were linked to a mixed type of DME, characterized by a combination of two or more types seen on OCT. Most patients presented with best corrected visual acuity ranging from 6/12 to 6/18. Furthermore, fasting, postprandial, and glycated hemoglobin levels did not show significant correlations with the type of DME on OCT, likely due to variations in treatment history and other health conditions. A strong correlation was observed between central macular thickness and the type of DME, with increased CMT associated with a higher occurrence of mixed DME. Among patients with proliferative diabetic retinopathy, mixed type DME was most commonly seen, while diffuse retinal thickening and cystoid macular edema were noted in patients with other forms of diabetic retinopathy.

## Data Availability

All data produced in the present study are available upon reasonable request to the authors

## TABLES

### RESULTS

**TABLE 1.** AGE WITH TYPE OF DME.

| DME | Diffuse<br>Retinal |  | Cystoid<br>type |  | Serous |  | VMT |  | Mixed |  |
| --- | --- | --- | --- | --- | --- | --- | --- | --- | --- | --- |
|  | RE | LE | RE | LE | RE | LE | RE | LE | RE | LE |
| 30 – 40<br>years | 1 | 0 | 2 | 1 | 0 | 0 | 0 | 0 | 1 | 0 |
| 40-50years | 2 | 0 | 3 | 3 | 1 | 0 | 1 | 0 | 1 | 2 |
| 50-60<br>years | 6 | 3 | 8 | 6 | 2 | 5 | 5 | 2 | 11 | 8 |
| 60 – 70<br>years | 9 | 8 | 14 | 5 | 0 | 0 | 4 | 6 | 9 | 5 |
| 70 – 80<br>years | 0 | 0 | 0 | 1 | 0 | 0 | 2 | 1 | 4 | 5 |
| Total | 18 | 11 | 27 | 18 | 3 | 5 | 12 | 9 | 2 | 20 |

**TABLE 2.** DESCRIPTION OF GENDER WITH TYPE OF DME.

| DME | Diffuse<br>Retinal |  | Cystoid<br>type |  | Serous |  | VMT |  | Mixed |  |
| --- | --- | --- | --- | --- | --- | --- | --- | --- | --- | --- |
|  | RE | LE | RE | LE | RE | LE | RE | LE | RE | LE |
| Male | 9 | 6 | 18 | 9 | 2 | 5 | 7 | 4 | 19 | 14 |
| Female | 9 | 5 | 9 | 7 | 1 | 0 | 5 | 5 | 7 | 6 |
| Total | 18 | 11 | 27 | 16 | 3 | 5 | 12 | 9 | 26 | 20 |

**TABLE – 3.**
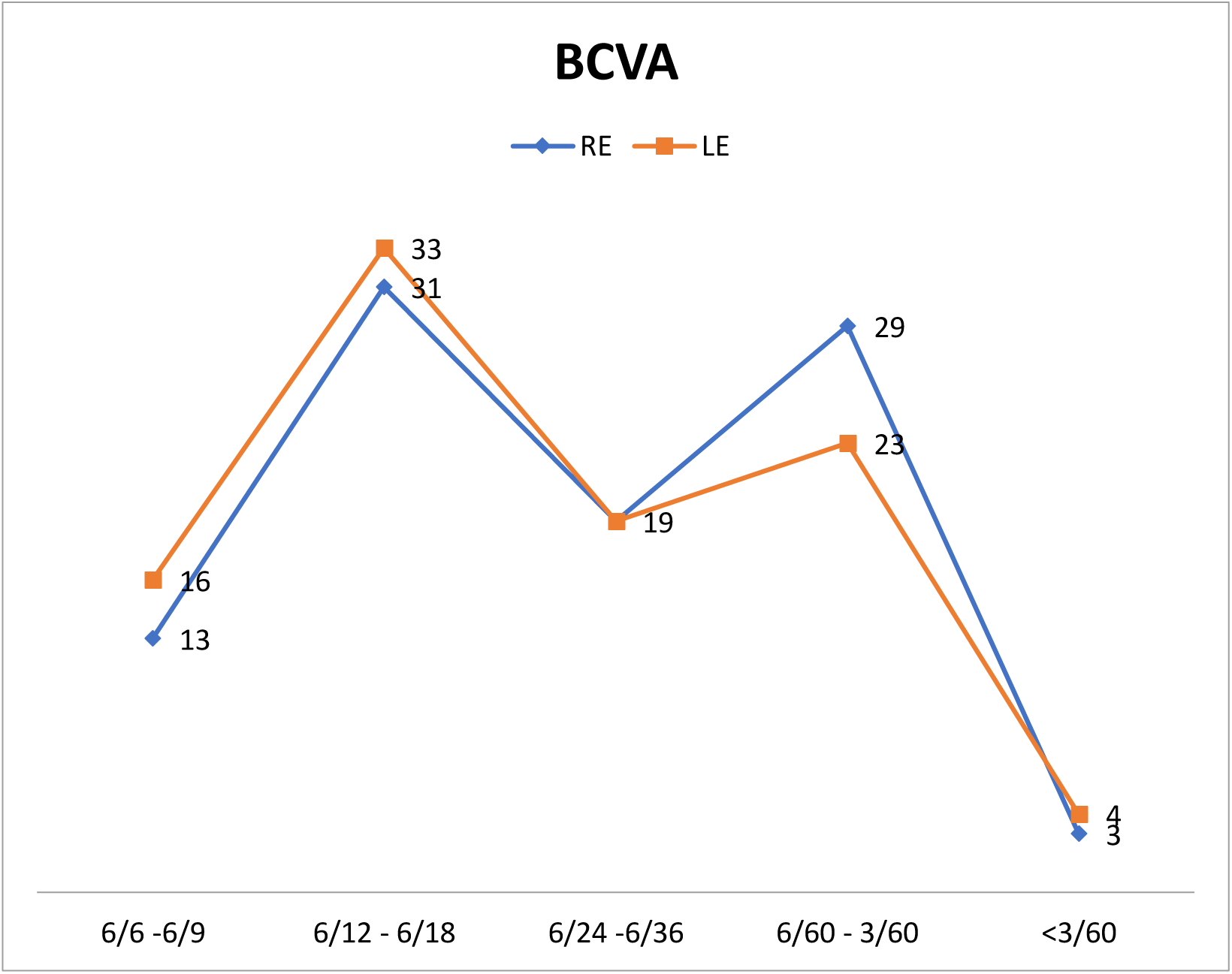
BEST CORRECTED VISUAL ACUITY ON PRESENTATION.

**TABLE – 4.**
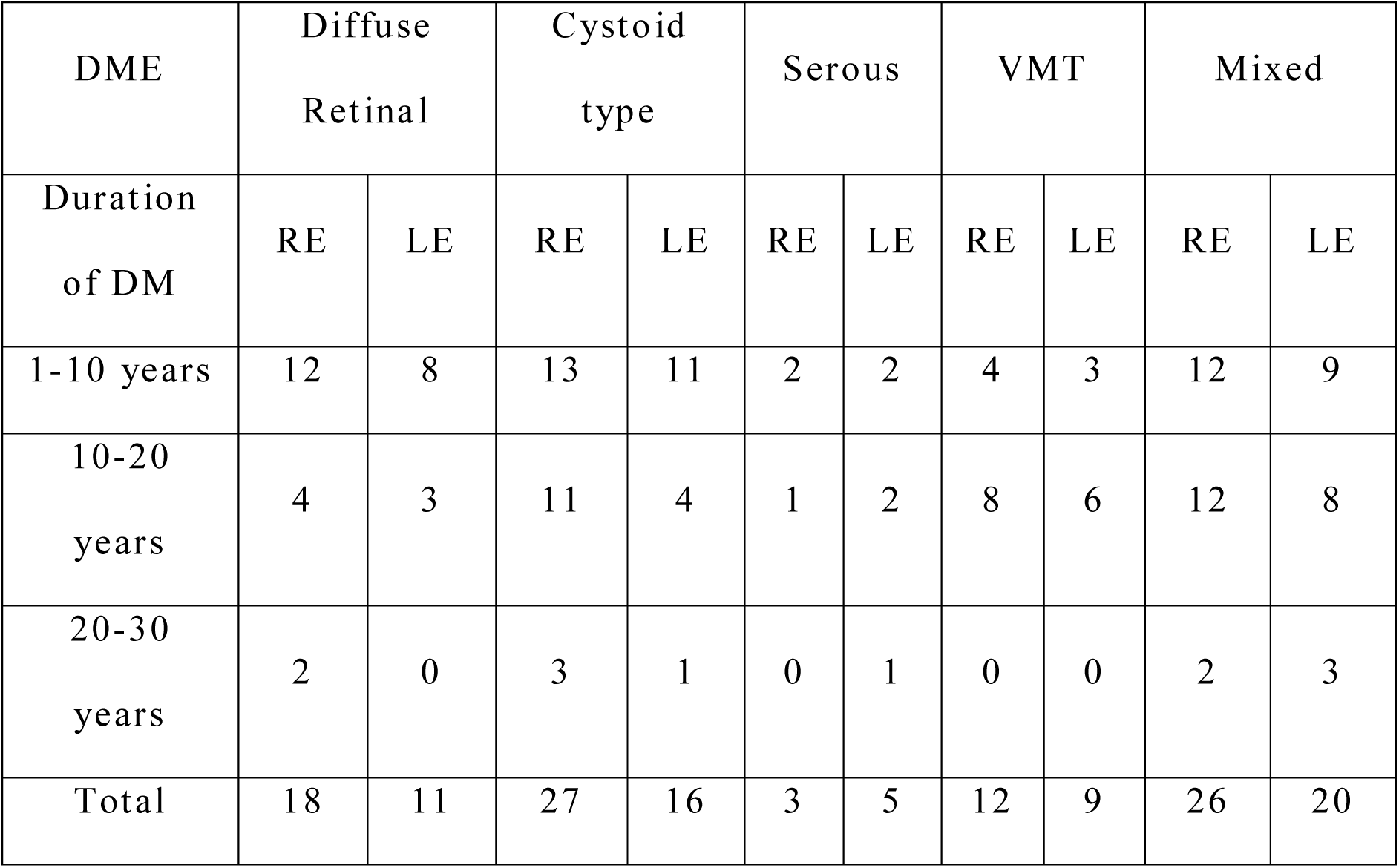
DURATION OF DM WITH TYPE OF DME ON OCT.

| DME | Diffuse Retinal |  | Cystoid type |  | Serous |  | VMT |  | Mixed |  |
| --- | --- | --- | --- | --- | --- | --- | --- | --- | --- | --- |
| Duration of DM | RE | LE | RE | LE | RE | LE | RE | LE | RE | LE |
| 1-10 years | 12 | 8 | 13 | 11 | 2 | 2 | 4 | 3 | 12 | 9 |
| 10-20 years | 4 | 3 | 11 | 4 | 1 | 2 | 8 | 6 | 12 | 8 |
| 20-30 years | 2 | 0 | 3 | 1 | 0 | 1 | 0 | 0 | 2 | 3 |
| Total | 18 | 11 | 27 | 16 | 3 | 5 | 12 | 9 | 26 | 20 |

**TABLE 5.** DIAGNOSIS OF DM WITH TYPE OF DME ON OCT.

| DME | Diffuse Retinal |  | Cystoid type |  | Serous |  | VMT |  | Mixed |  |
| --- | --- | --- | --- | --- | --- | --- | --- | --- | --- | --- |
| Diagnosis | RE | LE | RE | LE | RE | LE | RE | LE | RE | LE |
| Mild | 0 | 0 | 0 | 0 | 1 | 0 | 0 | 0 | 0 | 1 |
| Moderate | 6 | 4 | 5 | 0 | 0 | 2 | 1 | 2 | 2 | 5 |
| Sever | 6 | 5 | 12 | 7 | 1 | 2 | 2 | 0 | 7 | 2 |
| Very Severe | 0 | 0 | 0 | 1 | 0 | 0 | 1 | 0 | 1 | 0 |
| PDR | 6 | 2 | 10 | 8 | 1 | 1 | 8 | 7 | 16 | 12 |
| Total | 18 | 11 | 27 | 16 | 3 | 5 | 12 | 9 | 26 | 20 |

**TABLE 6.** FBS WITH TYPE OF DME ON OCT.

| DME | Diffuse<br>Retinal |  | Cystoid<br>type |  | Serous |  | VMT |  | Mixed |  |
| --- | --- | --- | --- | --- | --- | --- | --- | --- | --- | --- |
| FBS | RE | LE | RE | LE | RE | LE | RE | LE | RE | LE |
| Up to<br>100mg/dl | 6 | 3 | 8 | 4 | 2 | 0 | 2 | 4 | 8 | 5 |
| 101-200<br>mg/dl | 12 | 8 | 18 | 12 | 1 | 5 | 8 | 4 | 17 | 12 |
| 201-300mg/dl | 0 | 0 | 1 | 0 | 0 | 0 | 2 | 1 | 1 | 3 |
| Total | 18 | 11 | 27 | 16 | 3 | 5 | 12 | 9 | 26 | 20 |

**TABLE 7.**
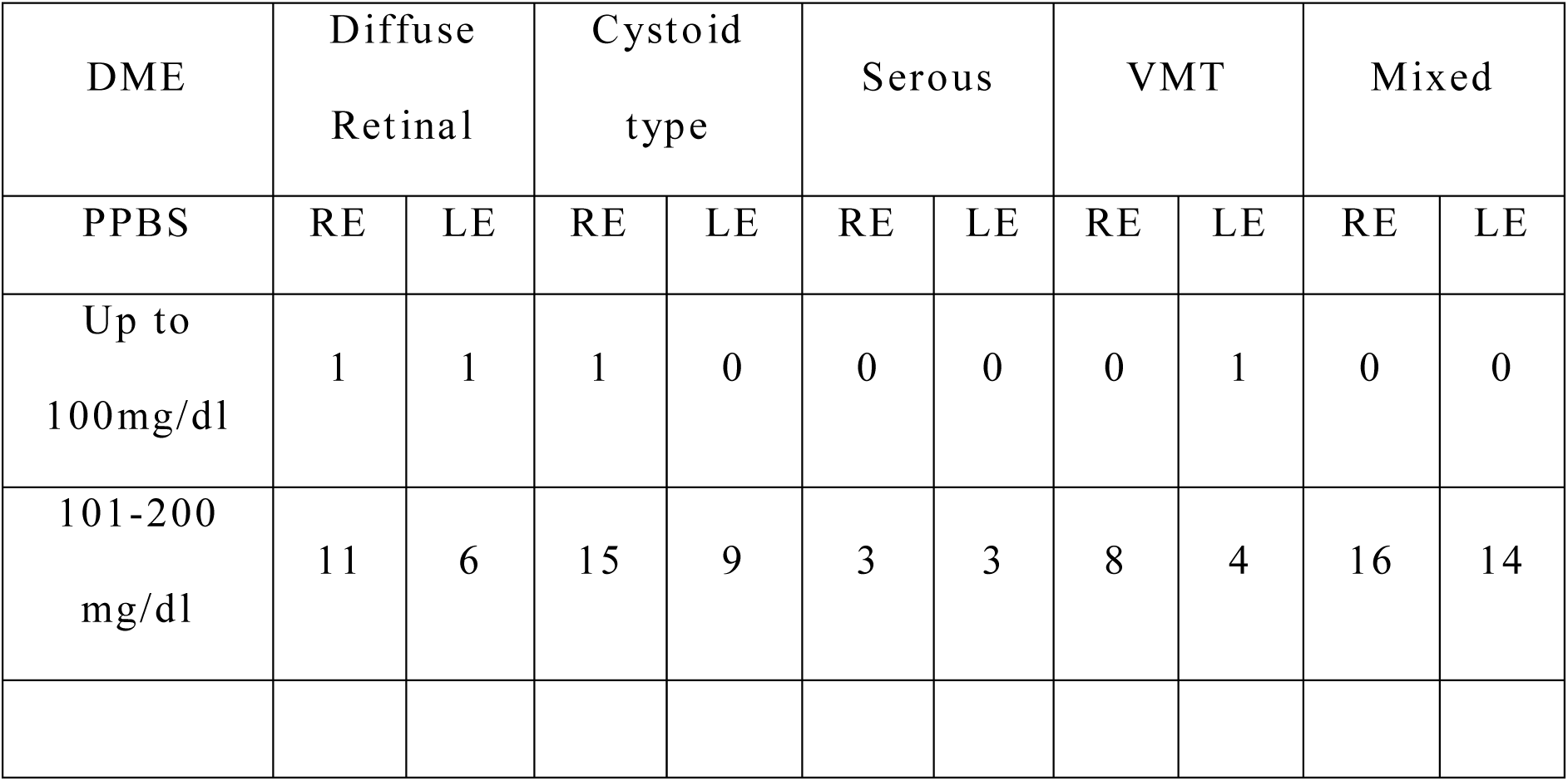
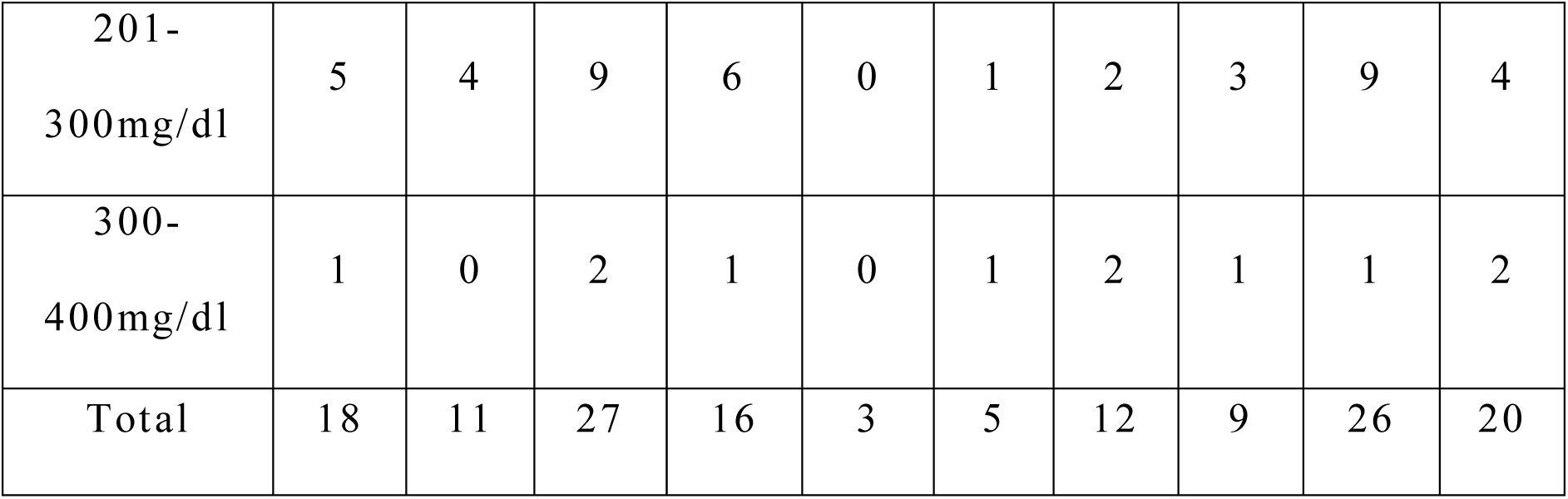
PPBS WITH TYPE OF DME ON OCT.

**TABLE 8.** CMT WITH TYPE OF DME ON OCT.

| DME | Diffuse<br>Retinal |  | Cystoid<br>type |  | Serous |  | VMT |  | Mixed |  |
| --- | --- | --- | --- | --- | --- | --- | --- | --- | --- | --- |
|  | RE | LE | RE | LE | RE | LE | RE | LE | RE | LE |
| 200-400<br>micron | 10 | 4 | 8 | 7 | 0 | 3 | 2 | 2 | 6 | 3 |
| 400-600<br>micron | 7 | 6 | 12 | 2 | 2 | 0 | 7 | 7 | 13 | 7 |
| 600-800<br>Micron | 0 | 1 | 6 | 2 | 0 | 1 | 2 | 0 | 4 | 3 |
| 800-1000<br>micron | 1 | 0 | 0 | 1 | 0 | 0 | 0 | 0 | 1 | 1 |
| 1000-1200<br>micron | 0 | 0 | 0 | 1 | 0 | 0 | 1 | 0 | 2 | 2 |
| Total | 18 | 11 | 26 | 12 | 2 | 4 | 12 | 7 | 26 | 16 |

## IMAGES

### Cystoid macular edema

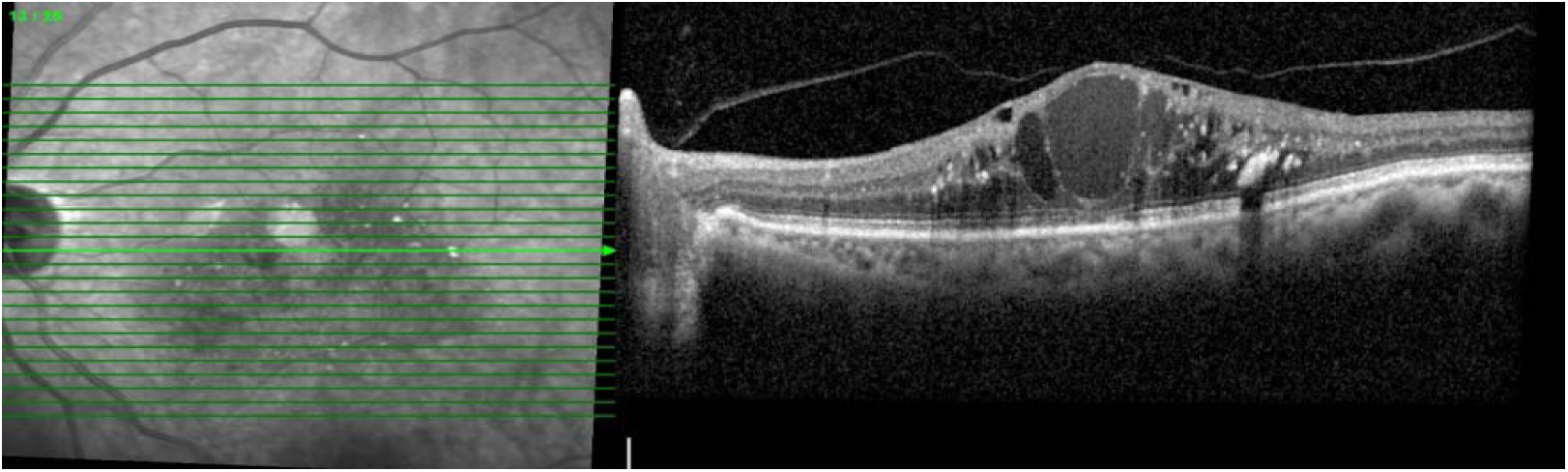

### Diffuse Spongiform retinal thickening

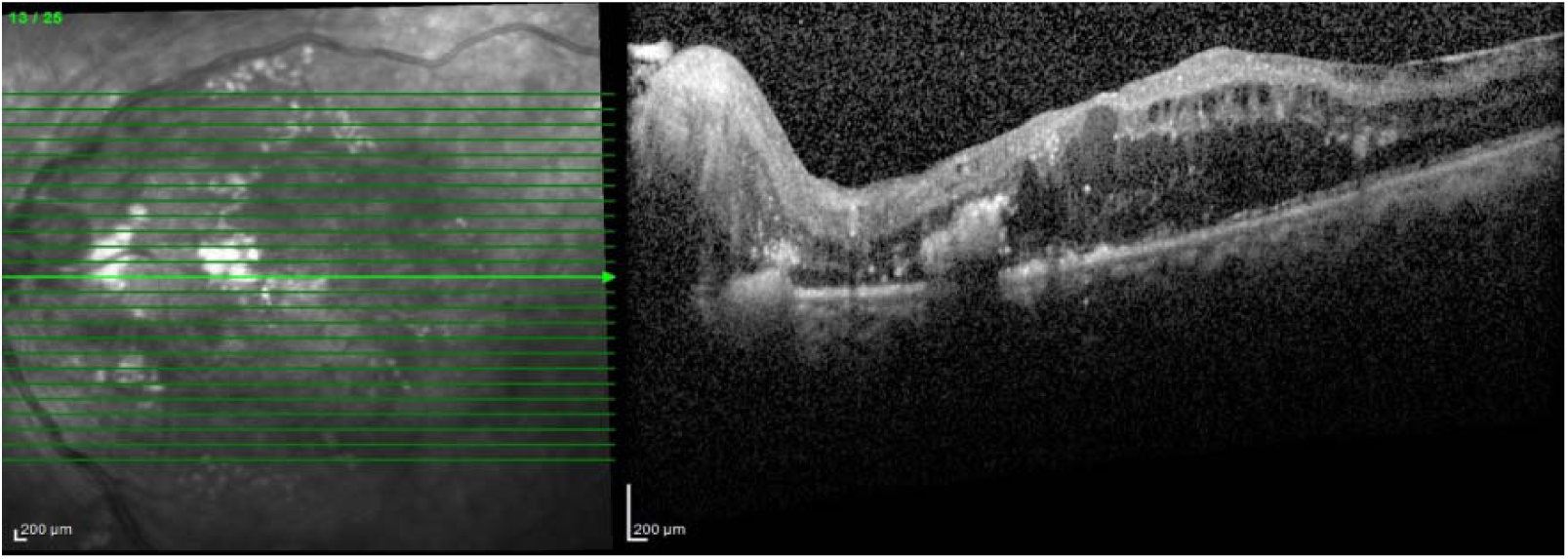

### Serous macular detachment

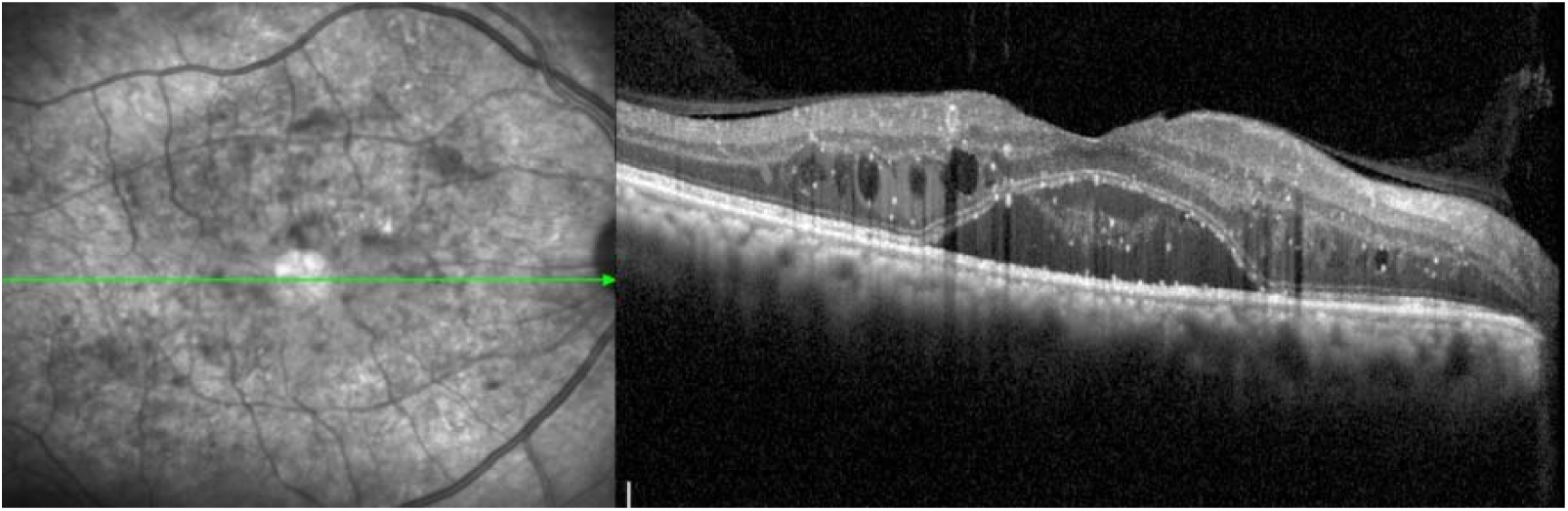

### Mixed type

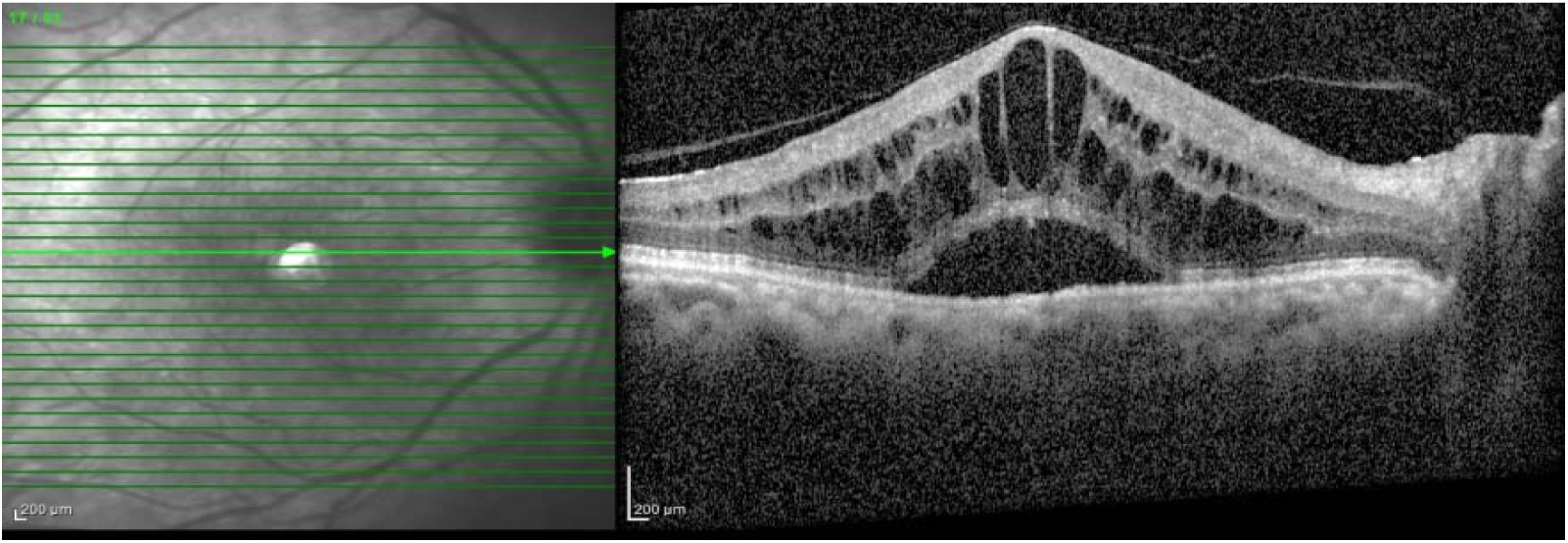

### Taut posterior hyaloid

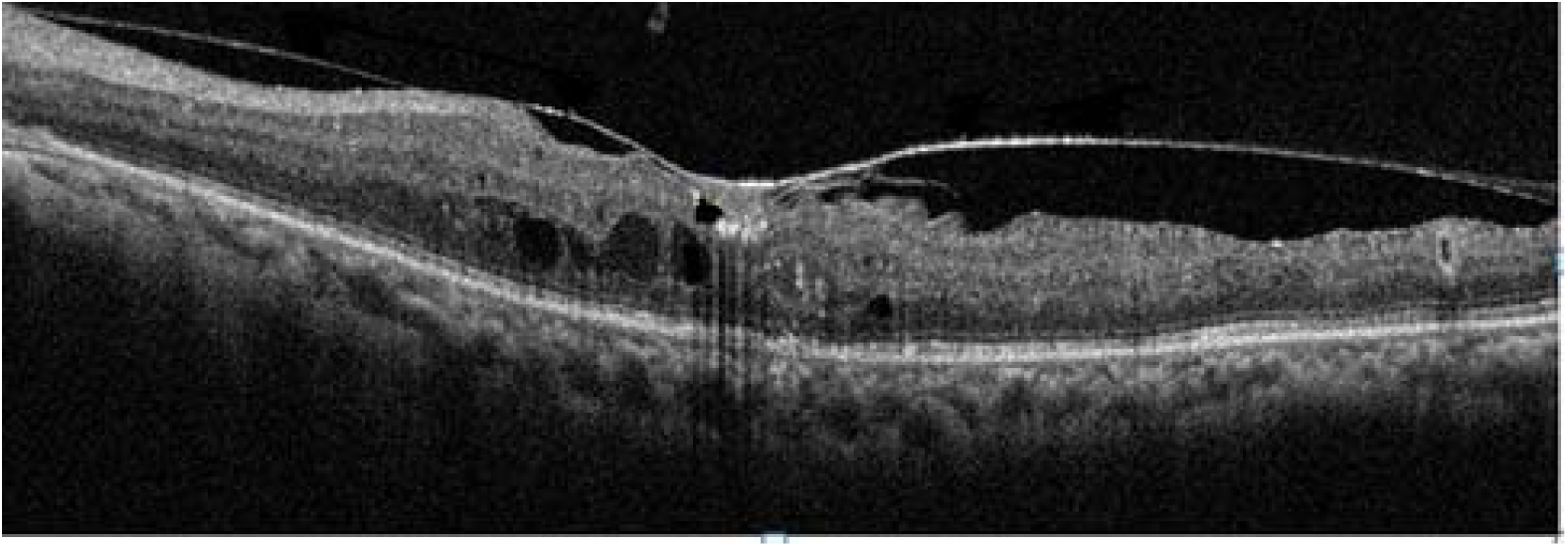

